# Electric Fan Use in Replicated 8-Hour Extreme Heat Event: Sex Differences in Systemic Biomarkers and Thermoregulation

**DOI:** 10.1101/2025.03.06.25323498

**Authors:** Faming Wang, Haojian Wang, Tze-Huan Lei, Huijuan Xu, Caiping Lu, Toby Mündel

## Abstract

Health agencies, including the Centers for Disease Control and Prevention (CDC) and World Health Organization (WHO), recommend different temperature thresholds for electric fan use during heat events (CDC: 32.2℃, WHO: 40℃). Nonetheless, these guidelines do not account for the fan’s physiological effects on immune and inflammatory responses in males and females. This study evaluated the efficacy of electric fan use in mitigating immune function, inflammation, and organ function during an eight-hour simulated extreme heat event replicating conditions in Hangzhou, China, on the August 3, 2024 (semi-hourly temperature and humidity fluctuations; average temperature: 39.9℃ [37.6-41.1℃], RH: 47.1% [40-57%]). Thirty-two young adults (16 males and 16 females) underwent three eight-hour trials: (1) no electric fan with limited fluid intake (500mL; Con), (2) fan use with limited fluid intake (500mL; Fan) and (3) fan use with sufficient fluid intake (3L; Fan+Fluid). Core temperature, cardiovascular responses, plasma electrolytes, stress hormones, inflammatory markers, and organ function biomarkers were assessed. Fan+Fluid significantly reduced core temperature, stress hormone levels, inflammatory responses, and organ function biomarkers in both sexes (all *p*<0.05). Notably, Fan+Fluid greatly reduced IL-6, IL-1β, ALT and BUN by 23.9%, 32.5%, 15.9% and 23.6%, respectively, compared to CON, despite the core temperature difference is marginal. Females exhibited consistently higher stress hormone levels, inflammatory responses, and organ function markers than males across all trials (all *p*<0.05). These findings highlight the benefits of electric fan use in prolonged extreme heat and suggest that females may require more intense cooling interventions due to heightened inflammatory and organ stress responses.

## INTRODUCTION

Prolonged heatwaves are an escalating global public health concern, exacerbating pre-existing health conditions and contributing to rising mortality rates.^1,2^ Heat-related illnesses, ranging from heat exhaustion to organ dysfunction and heatstroke, pose immediate risks during extreme temperature events.^3^ Vulnerable populations, including the elderly, children, and individuals with pre-existing cardiovascular, respiratory, or metabolic conditions, are particularly susceptible.^4^ Climate change, coupled with rapid urbanization, is projected to intensify the frequency, severity, and duration of extreme heat events,^5^ further straining public health systems and local healthcare infrastructures. Urban areas are disproportionately affected due to the urban heat island effect, characterized by dense populations, limited green spaces, and heat-absorbing infrastructure. These emerging challenges underscore the urgent need for robust, evidence-based strategies to mitigate the health impacts of heat stress, especially in regions where access to air conditioning and other advanced cooling technologies is limited.

Electric fans are often promoted as a low-carbon, cost-effective, and accessible cooling intervention during heatwaves; however, their efficacy and safety under extreme conditions remain contentious. Public health organizations, including the Centers for Disease Control and Prevention (CDC) and the World Health Organization (WHO), advise against fan use at temperatures exceeding 32.2–40°C, citing concerns that fans may exacerbate dehydration and increase heat strain.^6–8^ Similarly, the U.S. Environmental Protection Agency (EPA) discourages fan use in indoor environments above 32.2°C (90°F), recommending air conditioning or direct cooling methods instead.^9^ However, recent studies challenge these recommendations, demonstrating that fans can be effective under moderately high temperatures (up to 37–40°C) and relative humidity levels of 60%, especially for young adults.^10,11^ Evidence for older adults, however, remains more cautious, as fan use at extreme temperatures (≥35°C) and high humidity (≥70%) has been associated with increased cardiovascular strain and elevated core temperatures.^12^ These discrepancies highlight the need for more nuanced and context-specific recommendations for fan use, as fan performance is highly dependent on environmental conditions, humidity levels, and individual physiological factors.

A critical gap in the literature is the lack of consideration of sex differences in physiological responses during prolonged extreme heat exposure. Despite mounting evidence that males and females exhibit distinct thermoregulatory,^13^ cardiovascular, and hydration responses to heat,^14,15^ these differences remain underexplored in the context of electric fan use. This oversight has limited the development of tailored public health strategies that account for sex-specific vulnerabilities during extreme heat stress. Moreover, much of the existing research focuses narrowly on changes in core temperature (*T_core_*) during heat exposure. While *T_core_* is a key indicator of heat strain, it does not fully capture the systemic physiological effects induced by extreme heat.^16^ Heat stress can significantly affect systemic biomarkers of organ function, including indicators of kidney function,^17^ cardiovascular status, and inflammation,^18^ providing critical insights into the broader impacts of heat exposure. Addressing these gaps is essential to developing comprehensive and equitable public health recommendations.

The extreme heat event in Hangzhou, China, on August 3, 2024, exemplifies the urgent need for such research. On this day, temperatures exceeded 37°C for 8–9 consecutive hours (37.6–41.1°C), with high humidity levels (40–57%), resulting in a heat index of 50.1–54.7°C and wet-bulb temperatures of 29.1-30.7°C. Such prolonged exposure to extreme heat heightens the risk of heat-related illnesses and systemic organ dysfunction, particularly in urban areas where heatwave conditions are amplified. Existing studies on fan efficacy have largely employed short-duration (up to 3 hours), static exposure models under constant temperatures, failing to replicate the dynamic and prolonged nature of real-world heat events.^11,19–21^ Given the severity of heatwaves like the one in Hangzhou, understanding the full physiological impact, including sex-specific responses, is crucial for developing more effective cooling strategies.

We address these critical gaps by investigating the health impacts of electric fan use during a replicated extreme heatwave using a comprehensive framework. Unlike previous research, we employed a prolonged daily exposure model simulating fluctuating high temperatures and humidity over an extended period (8 hours), replicating the August 3rd Hangzhou heat event. This design enabled us to assess not only *T_core_* and cardiovascular responses but also systemic biomarkers of organ function and sex-specific physiological differences, providing a more holistic understanding of the health effects of fan use under extreme heat events. By broadening the scope of previous work, our findings aim to refine current public health guidelines and provide evidence-based insights into the risks and benefits of fan use during prolonged extreme heat events.

## METHODOLOGY

### Ethical approval and Participants

Ethical approval was secured from the Institutional Review Board of Xi’an University of Science and Technology (XUST-IRB224002), and the study was registered with the Chinese Clinical Trial Registry (ChiCTR2300071885). Participants provided verbal and written informed consent prior to enrollment, in strict accordance with the Declaration of Helsinki. A total of 32 healthy young adults were recruited: 16 males (age: 24.1±1.6 yr, body surface area: 1.75±0.04 m^2^; body mass index [BMI]: 22.2±1.6 kg/m^2^) and 16 females (age: 22.1±1.7 yr, body surface area: 1.61±0.06 m^2^; BMI: 20.1±1.83 kg/m^2^). To minimize hormonal variability, female participants were tested during the early follicular phase of their menstrual cycle, characterized by low progesterone levels.^22^ Anthropometric characteristics were not matched between participants to preserve external validity.

### Rationale for studying young adults

The inclusion of young males and females in this study was guided by two key considerations. While older adults are more susceptible to classical heatstroke during heatwaves due to diminished thermoregulatory capacity, the health risks faced by younger individuals under prolonged humid heat exposure remain less understood. Younger adults, particularly those engaged in outdoor occupations such as agriculture, construction, and manual labor, experience extended periods of extreme heat, posing unique health challenges.^23^ Unlike older populations, they are more likely to accumulate heat stress over time without immediate signs of classical heatstroke. Additionally, their engagement in physical activities or occupational tasks increases metabolic heat production, further compounding their vulnerability to heat-related illnesses. The absence of comprehensive data on how prolonged and repeated heat exposure affects this demographic underscores the need to examine their physiological responses under extreme conditions.

Beyond thermoregulatory failure, extreme humid heat exposure can trigger systemic effects, including early inflammatory responses. This study focuses on inflammation as a precursor to heat-related illnesses, such as systemic inflammatory response syndrome (SIRS), which may progress to heatstroke if heat exposure persists.^18^ By analyzing inflammatory markers, particularly interleukin-6 (IL-6), following an 8-hour heat exposure, this research seeks to identify early physiological indicators of heat stress that are often overlooked in younger populations. Understanding these early responses is critical not only for recognizing the susceptibility of younger individuals to extreme humid heat but also for informing public health strategies to mitigate risks. Given the increasing frequency and intensity of heat events due to climate change, addressing these gaps is essential for developing targeted interventions to prevent heat-related illnesses in younger, active populations working or exercising in extreme heat environments.

### Justification for the prolonged heat exposure

The August 3rd, 2024, heatwave in Hangzhou (eastern China, longitude: 120.1552° E; latitude: 30.2741° N) exemplifies the increasing frequency and severity of extreme humid heat events.^24^ To contextualize this event within a broader global framework, Raymond et al. analyzed the highest recorded extreme wet-bulb temperatures (*T_W_*) using the HadISD dataset, covering weather station data from 1979 to 2017.^25^ Their findings indicate that regions such as South Asia, parts of the Middle East, and Southeast Asia are particularly prone to recurrent extreme humid heat events, with Hangzhou emerging as a significant hotspot. The August 2024 heatwave was one of the most severe recorded in the region over the past two decades, underscoring the escalating risks driven by climate change.

Climate models project that by 2100, tropical and subtropical regions of Asia, particularly the Ganges and Indus River basins, will experience 6-hour wet-bulb temperatures exceeding 30°C, with some surpassing the critical 35°C threshold considered near or beyond human survivability.^26^ These extreme conditions will disproportionately affect densely populated agricultural regions in southern Asia, where prolonged heat exposure poses severe risks. The increasing frequency of such events is partly attributed to moisture transport from the warming Indian Ocean, contributing to rising atmospheric humidity levels. In this context, the August 2024 Hangzhou heatwave serves as a striking example of the intensifying threat posed by climate change. To enhance the ecological validity of our study, we replicated the diurnal fluctuations in temperature and relative humidity observed during this heatwave. **Figure 1** illustrates the semi-hourly fluctuations in environmental conditions during our 8-hour simulated heat event.

**Figure 1.**
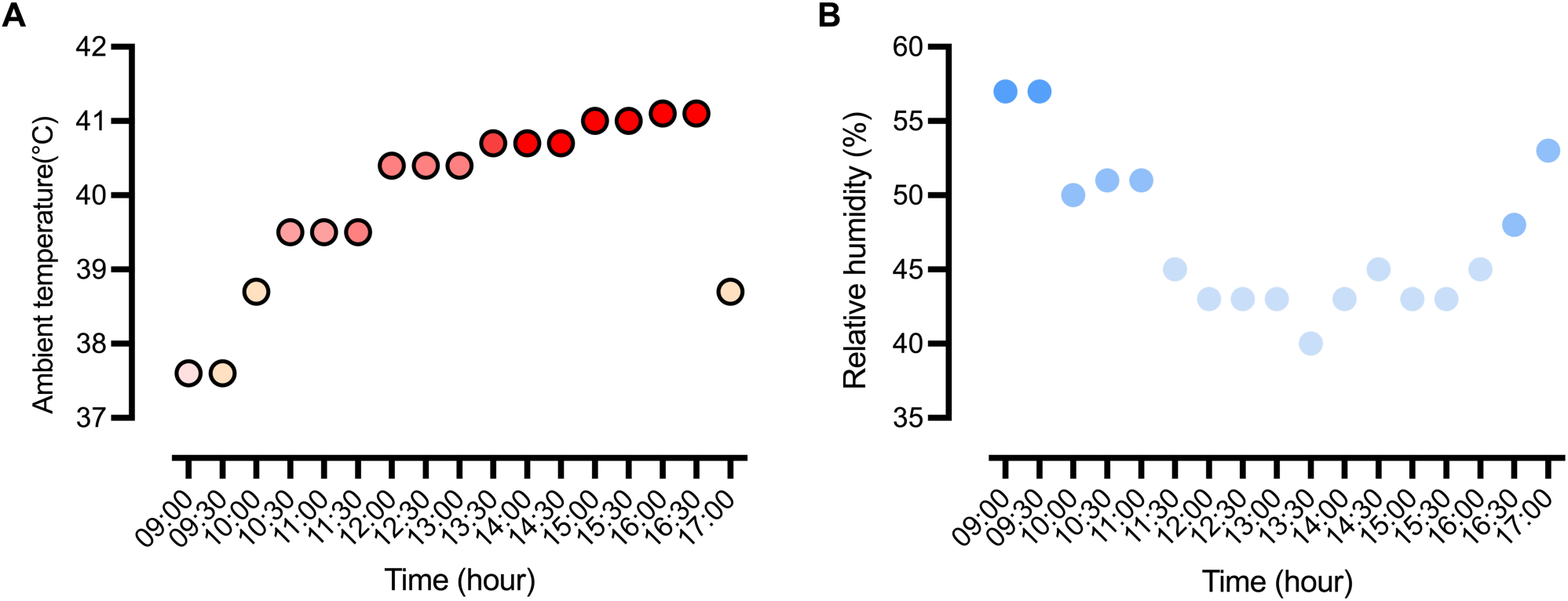
Semi-hourly fluctuations of ambient temperature and relative humidity during the 8-hour simulated heat wave event. Panel **A** shows the progressive increase in temperature, represented by a gradient shift from light to dark red, corresponding to the heat event’s duration. Panel **B** illustrates the gradual decrease in relative humidity, indicated by a gradient shift from dark to light blue. Together, these panels demonstrate the realistic diurnal variations in environmental conditions, reflecting the dynamic interplay between rising temperatures and falling humidity over the course of the 8-hour simulation.

### Experimental overview

Participants underwent three randomized and counterbalanced 8-hour heat event trials: Control (Con), electric fan use (Fan), and electric fan with 3L fluid consumption (Fan+Fluid). The temperature and humidity during these trials were designed to replicate the heat event reported in Hangzhou, China, on August 3, 2024, with an average temperature of 39.9℃ and 47.1% RH (relative humidity), and a wet-bulb temperature (*T_W_*) of 30.0℃(*T_W_*=29.1-30.7°C). During the Con and Fan trials, participants were restricted to consuming 500 mL of an electrolytes drink formulated with 10.8 mg of zinc, 0.5 g of carbohydrates, 283 mg of sodium, and isotonic properties. In the Fan+Fluid trials, participants were provided with 3 L of the same electrolyte drink. The fluid volumes were determined based on pilot study results, which revealed that consuming 500 mL maintained a moderate level of dehydration (less than 4% nude body weight loss), while 3 L maintained euhydration status by the end of the 8-hour heat exposure.

Trials were separated by 5-day washout periods to ensure baseline recovery of systemic inflammation markers. All trials were conducted between April and June in Xi’an, China (longitude: 108.9541° E; latitude: 34.2658° N). Each trial ran from 9 a.m. to 5 p.m. Before each trial, participants provided urine samples to confirm a urine-specific gravity of below 1.020.^27^ If the specific gravity exceeded this threshold, an isotonic electrolyte drink was administered until compliance was achieved. Pre-trial measurements included nude body weight and venous blood sampling. Participants were then seated in office chairs within a chamber (Espec Corp., Osaka, Japan), which was programmed to simulate the specified heat conditions. Participants engaged in routine sedentary activities such as reading and computer work, with an energy expenditure of approximately 1.0-1.4 mets (**Table 2**). They wore short-sleeve T-shirts, sports bras, underwear, trousers, socks, and shoes, resulting in total clothing insulation values of 0.30 clo for males and 0.31 clo for females. Dietary intake was standardized by providing participants with a BigMac burger (550 kcal) during each trial. Post-trial venous blood samples were collected 40–60 minutes after the eight-hour prolonged heat exposure to capture the peak levels of systemic biomarkers, allowing for a more accurate assessment of physiological strain induced by heat stress.^28^

### Electric fan use

An electric tower fan (Changhong model 73298401, Sichuan Changhong Electric Co. Ltd., Mianyang, China) was positioned 60 cm from each participant. Participants self-selected fan use based on their perceptions and adjusted air speeds: low (averaged speed: 1.94 m/s), moderate (2.17 m/s) and high (2.43 m/s). The average air speed was measured using a hot-wire anemometer (TES-1341N, TES Electrical Electronics Corp., Taipei, Taiwan), positioned 60 cm from the fan vents at heights of 0.1 m, 0.7 m, and 1.1 m. Detailed records of fan use, including start time, selected air speed, and total fan usage time at each air speed, were maintained (see **Table S1** in the Supplemental Material).

### Biochemical markers

#### Serum analysis

Venous blood samples were collected into clot activator tube (Additive-free vacuum blood collection tubes, KWS Medical Technology Co. Ltd., Shijiazhuang, China), allowed to clot, and centrifuged at 4°C, 3,000 × g for 20 minutes. The resulting serum was aliquoted into Eppendorf tubes (2 mL centrifuge tubes, BKMAM, Changde, China) and stored at -80 ℃. Serum analysis was performed using enzyme-linked immunosorbent assays (ELISAs). Inflammatory cytokines, including interlukin-6 (EK106HS-02, 4.7% intra-assay CV, sensitivity 0.02 pg/mL), interlukin-1β (EK101BHS -01, 4.5% intra-assay CV, sensitivity 0.02 pg/mL), and interferon-γ (EK180HS-AW1, 4.3% intra-assay CV, sensitivity 0.04 pg/mL), were measured using kits from Lianke Biotech Co. (Hangzhou, China). Intestinal permeability was assessed using intestinal fatty acid-binding protein kits (IFABP, SEA559Hu, 10% intra-assay CV, sensitivity 0.062 ng/mL, Youersheng, Wuhan, China), and cortisol levels were determined using ELISA kits (HEA462Ge, 10% intra-assay CV, sensitivity 0.56 ng/mL, Youersheng, Wuhan, China). Biomarkers for organ function, including blood urea nitrogen (BUN) and alanine transaminase (ALT), were analyzed using an enzyme label detector (Epoch, BioTek, Winooski, USA) and an automatic biochemistry analyzer (Chemray 800, Radu Life Sciences, Shenzhen, China), respectively.

#### Plasma analysis

Plasma was separated from whole blood samples collected into EDTA-coated tubes (heparin tubes, KWS Medical Technology Co. Ltd., Shijiazhuang, China) by centrifugation at 4 °C, 3,000 × g for 15 minutes. Plasma electrolytes (Na⁺, K⁺, Cl⁻) were measured in triplicate using an automatic ion-selective electrode analyzer (Biobase BKE-C, Biobase Bioindustry Co. Ltd., Ji’nan, China).

#### Capillary blood analysis

Capillary blood samples were collected pre- and post-exposure from participants in a seated position after a minimum of 15 minutes of rest. Leukocyte count, hemoglobin, and hematocrit were analyzed using an automatic hematology analyzer (Getein BHA-3000, Biotech Inc., Nanjing, China). All results were corrected for plasma volume changes.^29^

### Body temperature, cardiovascular and metabolic measurements

Core temperature (*T*_core_) was continuously monitored using a rectal thermistor (YSI401, Yellow Spring Instrument, Yellow Springs, OH; accurate: ±0.1°C) inserted 10 cm beyond the anal sphincter. Mean skin temperature (*T*_sk_) was calculated using readings from four iButton dataloggers (model number DS1992L, Maxim Integrated Product Inc., San Jose, CA) positioned at standard sites and averaged using the Ramanathan’s formula.^15^ Heart rate was continuously recorded using a Polar Vantage XL monitor (Polar Electro Oy, Kempele, Finland). Brachial artery blood pressure was measured at baseline and post-exposure using an automatic blood pressure monitor (YE660CR, Yuwell Medical Instruments Co. Ltd., Yancheng, China). Mean arterial pressure (MAP) was calculated using the following equation:

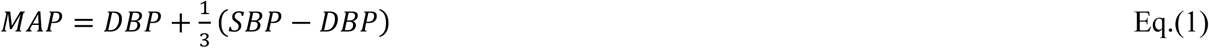

where DBP and SBP are diastolic and systolic blood pressures, respectively.

The metabolic rate was obtained using a portable metabolic system (COSMED K5, COSMED Srl, Rome, Italy). The system was calibrated before each trial using zero and β-standard gas concentrations (CO_2_: 5%, O_2_: 16%, with nitrogen balanced) and a 3-liter syringe (Hans Rudolph 3L Calibration Syringe, FIM Medical, Lyon, France).

### Perceptions and psychological stress

Thermal comfort (7-point scale, -3: very uncomfortable to +3: very comfortable,^30^ thermal sensation (9-point scale, -4: extremely cold to +4: extremely hot,^31^ thirst sensation (7-point scale, 1: not thirsty at all to 7: very, very thirsty,^32^ and psychological stress (10-point scale, 0: no psychological stress to 10: very severe psychological stress)^33^ were surveyed at 30-minute intervals throughout the 8-hour heat exposure.

### Hydration status

The dehydration rate was calculated by accounting for sweat production, fluid intake, and urine loss using the following equation:

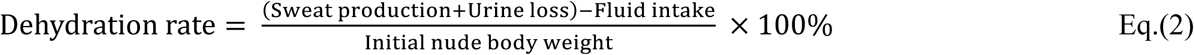

### Statistical analysis

All statistical analyses were performed with SPSS software for Windows (IBM SPSS Statistics 20, NY, USA) and figures were created using GraphPad Prism (version 7.00, Insight Venture Management LLC, New York, USA). Homogeneity of variance was examined by Levene’s test, and data normality was assessed by the Kolmogorov-Smirnov Test. The effects of different conditions (Con, Fan, Fan+Fluid) on body temperature, cardiovascular and inflammatory responses, and organ function biomarkers across both sexes and timepoints were analyzed using a two-way mixed-model ANOVA. Where main or interaction effects occurred, post hoc pairwise analyses were performed using the Bonferroni correction method. Data are presented as means (SD), unless otherwise stated. Statistical significance was set at *p*≤0.05.

## RESULTS

### Core temperature and cardiovascular responses

Core temperature did not differ between sexes (**Figure 2**, *p* = 0.97) but showed a significant interaction between conditions and timepoints (*p*<0.01). Specifically, core temperature in the Fan+Fluid trial was consistently lower than in the Con and Fan trials from the 4-hour mark onwards for both sexes. Systolic blood pressure (SBP), diastolic blood pressure (DBP), and mean arterial pressure (MAP)were significantly different between sexes (*p*<0.05). Males exhibited higher SBP and MAP compared to females at baseline and at the end of the 8-hour heat event, while DBP was higher in males than females only at the end of the heat exposure (all *p*<0.018, **Table 2**). No interaction between conditions and timepoints was observed for SBP, DBP, or MAP (all *p*>0.13). However, SBP and MAP were consistenly lower in females compared to males acorss all conditions (Con, Fan and Fan+Fluid) at the conclusion of the 8-hour heat event. Heart rate did not differ between sexes but revealed a significant interaction effect between conditions and timepoints (*p*=0.011).Specifically, heart rate was lower in the Fan+Fluid condition compared to Con, but this reduction was observed only in male participants.

**Figure 2.**
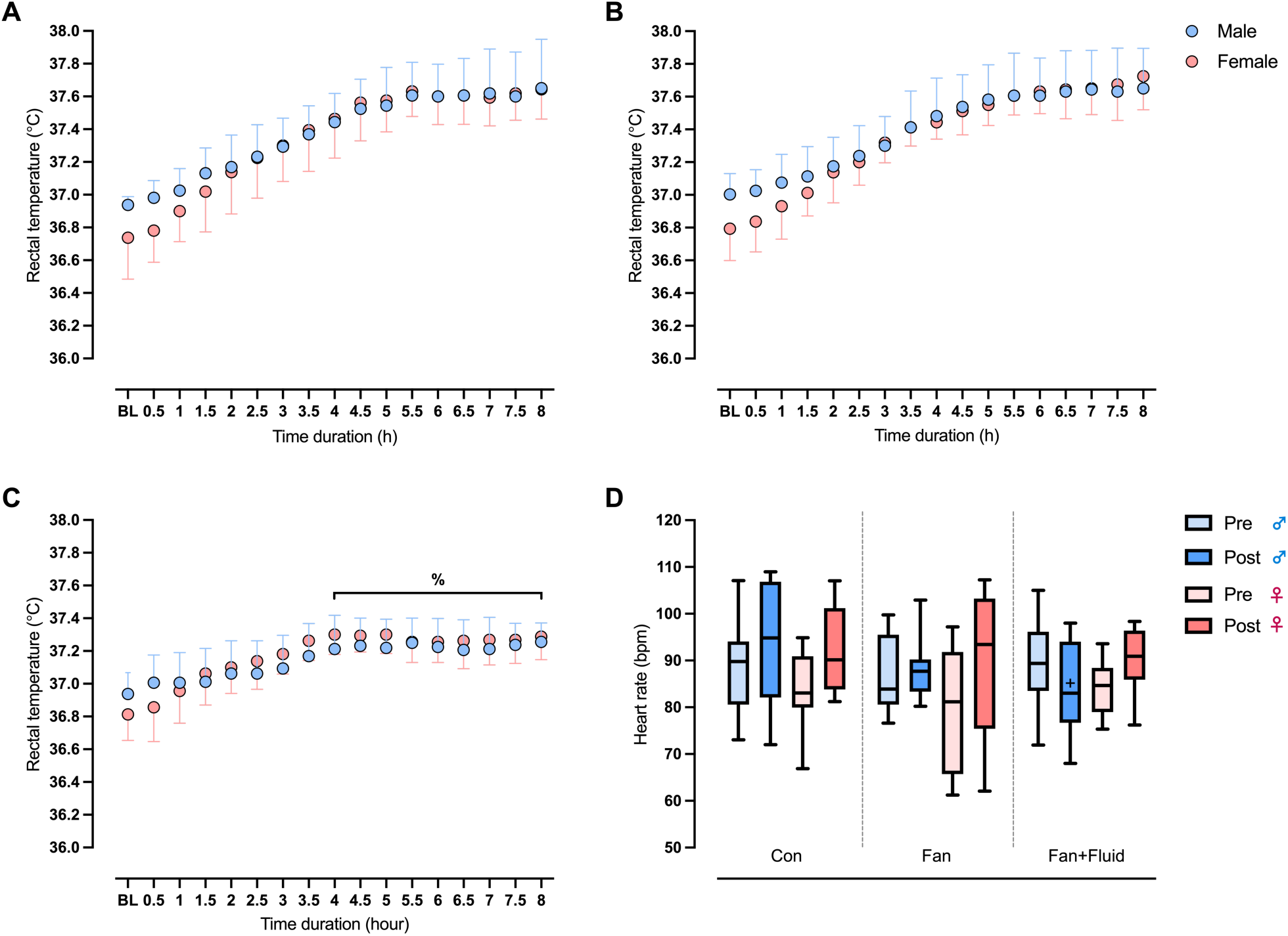
Semi-hourly core temperature (*T_core_*) response at baseline (BL) and throughout the eight-hour exposure for both sexes (A, Con; B, Fan; C, Fan+Fluid) as well as heart rate responses (D) at pre-exposure (pre) and at the end of the exposure (post) for both sexes. %, indicates *T_core_* in Fan+Fluid was significantly different from that in Con and Fan for both sexes (*p*<0.05); +, Fan+Fluid significantly different from Con at the corresponding time point (*p*<0.05).

### Hydration status

Dehydration rate was significantly greater in females than males during the Con and Fan trials (**Table 1**, *p*<0.01). Consequently, plasma volume reduction was greater in females compared to males across all conditions (**Figure 3**, *p*<0.01). Electrolyte balance between sexes showed limited differences, with chloride ion (Cl^-^) concentration being higher in females than males at the end of the Con trial (*p*<0.05). Plasma electrolytes revealed an interactional effect between conditions and timepoints (all *p*<0.05). Sodium concentrations were lower in the Fan+Fluid condition compared to both Con and Fan for both sexes (all *p*<0.05) Potassium (K^+^) concentrations were higher in the Fan condition compared to Fan+Fluid, but this effect was observed only in female participants. For chloride (Cl^-^) concentrations, males exhibited higher levels in the Fan condition than in Fan+Fluid, whereas females had lower chloride concentrations in Fan+Fluid compared to both Con and Fan at the end of the 8-hour heat event.

**Figure 3.**
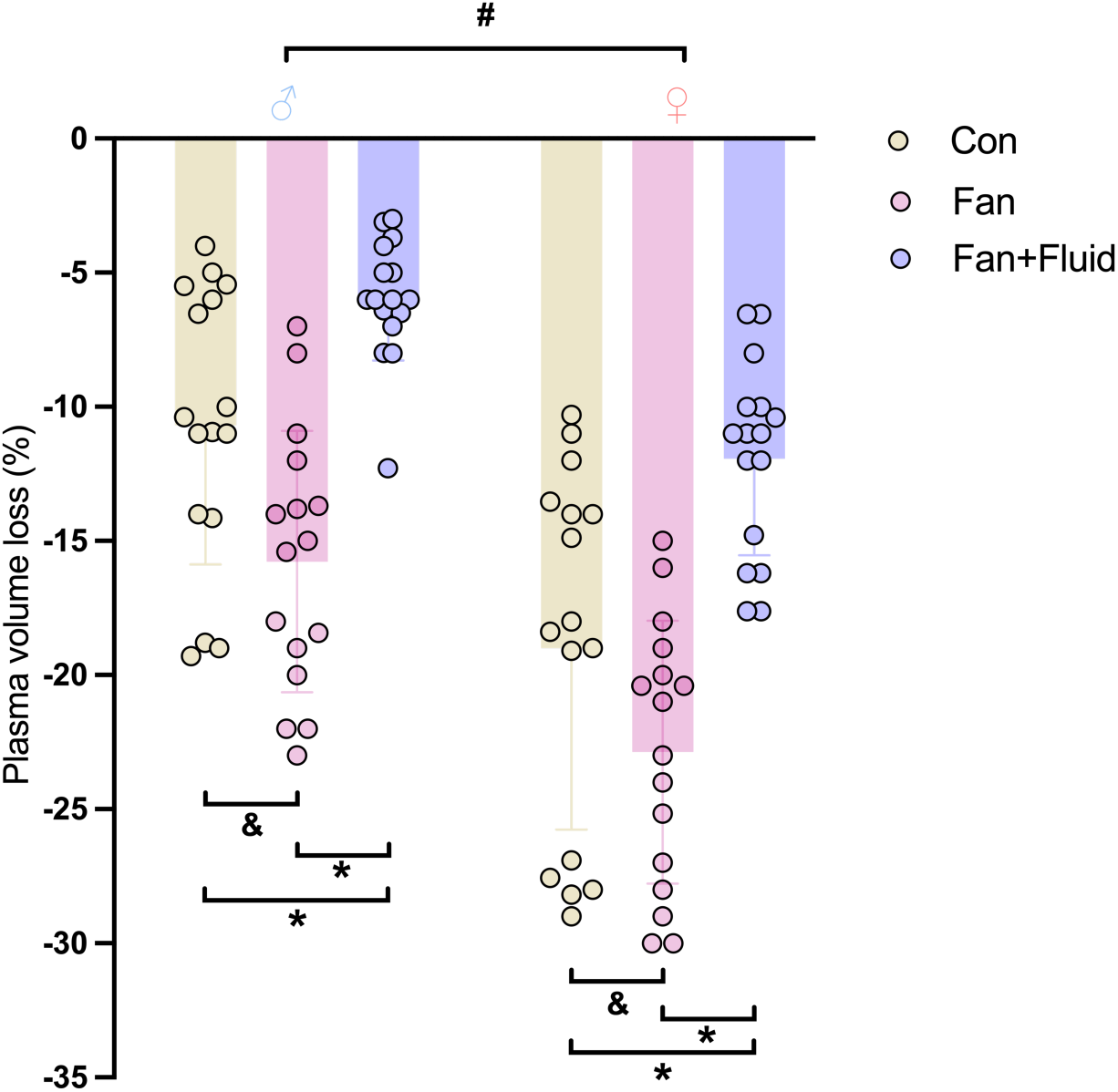
Plasma volume (PV) loss from baseline across different conditions (Con, Fan and Fan+Fluid) for both sexes. &, indicates a significant difference between Con and Fan (*p*<0.05); #, indicates significant differences between sexes across all experimental conditions (*p*<0.05); *, indicates significant differences between Fan+Fluid and Con or Fan (*p*<0.05).

**Table 1.**
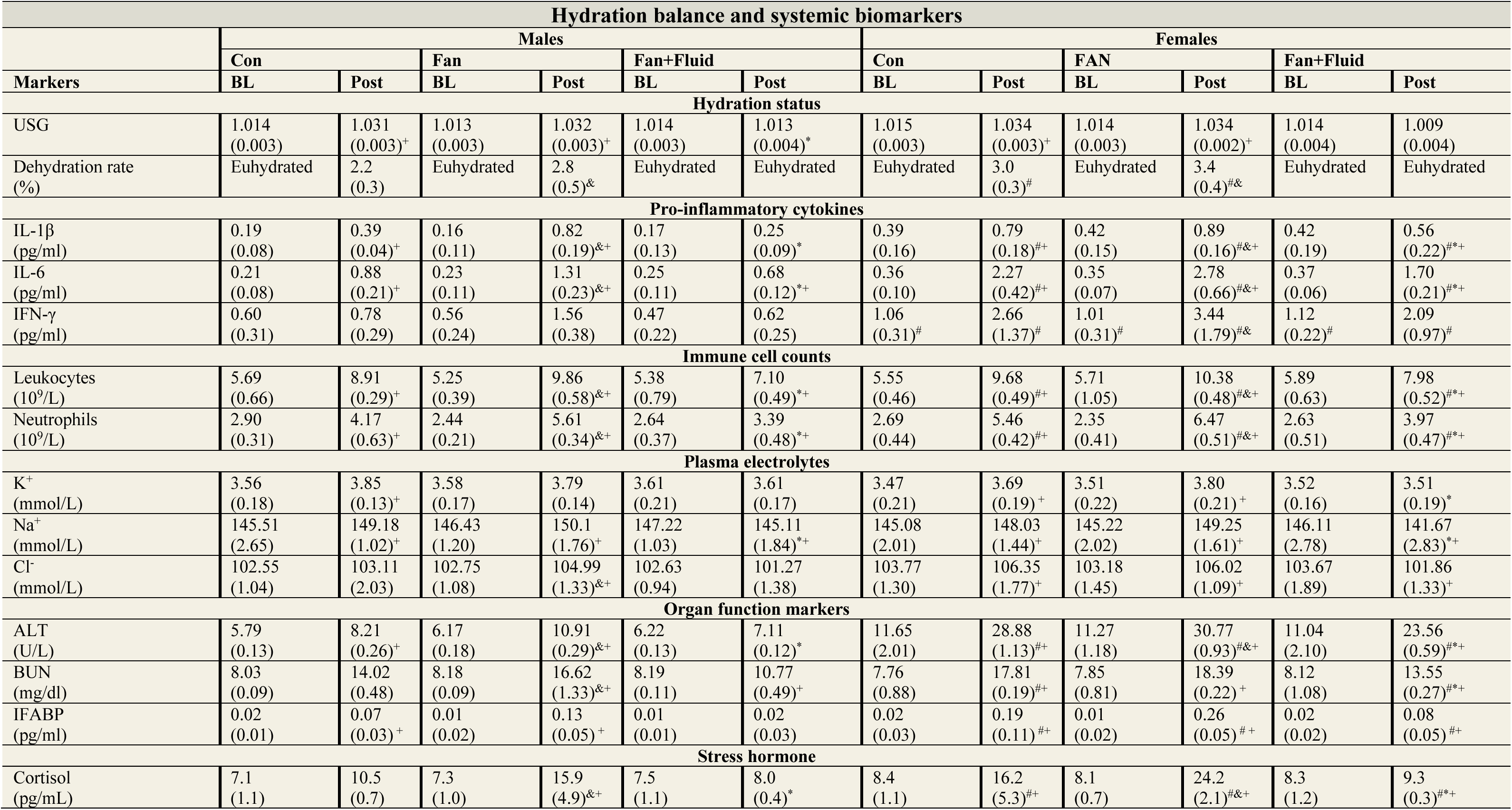
Hydration status and systemic biomarkers at baseline (BL) and at the end of the 8-hour replicated heat exposure (Post) across three trial conditions (Con, Fan, and Fan+Fluid). -, not applicable; +, indicates a significant difference than Pre; &, indicates a significant difference between Con and Fan (*p*<0.05); #, indicates significant differences between sexes within the corresponding trial condition (*p*<0.05); *, indicates significant differences between Fan+Fluid and Con or Fan (*p*<0.05). The numbers in parentheses represent standard deviation (SD).

### Inflammatory responses and immune cell count

Interleukin-6 (IL-6), interleukin-1β (IL-1β), interferon-γ (IFN-γ), leukocyte and neutrophil counts significantly differed between groups (**Table 1**, all *p*<0.01). Interaction effects between conditions and timepoints were observed for IL-6, IL-1β, leukocyte, and neutrophil counts (all *p*<0.01), but not for IFN-γ. Females exhibited consistently higher concentrations of IL-6 (Con: 257.9% higher; Fan: 212.2% higher; Fan+Fluid: 250% higher), IL-1β (Con: 202.6% higher; Fan: 108.5% higher; Fan+Fluid: 224% higher), IFN-γ (Con: 341% higher; Fan: 220.5% higher; Fan+Fluid: 337.1% higher), leukocytes (Con: 8.6% higher; Fan: 5.3% higher; Fan+Fluid: 12.4% higher), and neutrophils (Con: 30.9% higher; Fan: 15.3% higher; Fan+Fluid: 17.1% higher) compared to males at the end of the 8-hour heat event across all conditions. Among the interventions, the Fan+Fluid condition resulted in the lowest concentrations of IL-6, IL-1β, leukocytes, and neutrophils compared to Con (IL-6: -23.9%; IL-1β: - 32.5%; leukocytes: -18.9%; neutrophils: -23.0%) and Fan (IL-6: -43.5%; IL-1β: -53.3%; leukocytes: - 25.6%; neutrophils: -39.1%) at the end of the 8-hour heat event for both sexes (all *p*<0.01). Conversely, the Fan condition resulted in the highest concentrations of these biomarkers compared to both Con (IL-6: 35.7% higher; IL-1β: 61.5% higher; leukocytes: 9.0% higher; neutrophils: 26.5% higher) and Fan+Fluid (IL-6: 78.1% higher; IL-1β: 143.5% higher; leukocytes: 34.5% higher; neutrophils: 64.2%) for both sexes at the end of the 8-hour heat event.

### Organ function markers

Alanine transaminase (ALT), blood urea nitrogen (BUN), and intestinal fatty acid-binding protein (IFABP) differed significantly between groups (**Table 1**, *p*<0.01) and revealed interactional effects between conditions and timepoints (*p*<0.01). Specifically, ALT (Con: 351.8% higher; Fan: 282.0% higher; Fan+Fluid: 331.4% higher), BUN (Con: 127.0% higher; Fan: 110.7% higher; Fan+Fluid: 125.8% higher), IFABP (Con: 271.4% higher; Fan: 200% higher; Fan+Fluid: 400% higher) concentrations were higher in females compared to male across all conditions. The Fan+Fluid intervention yielded the lowest concentrations of ALT, BUN, and IFABP in both sexes at the end of the 8-hour heat event, outperforming both Con (ALT: 15.9% lower; BUN: 23.6% lower; IFABP: 64.7% lower) and Fan (ALT: 29.1% lower; BUN: 30.8% lower; IFABP: 76.9% lower). Conversely, the Fan condition resulted in higher ALT levels in both sexes compared to Con (19.7% higher) and Fan+Fluid (42.0% higher), while BUN increased only in males relative to Con (18.5% higher) at the trial’s end. In comparison to Fan+Fluid, the Fan condition also elevated BUN markedly, with increases of 54.3% in males and 35.7% in females by the conclusion of the 8-hour heat event.

### Stress hormone

The cortisol response differed between groups (*p*<0.01) and revealed an interactional effect between conditions and timepoints (*p*<0.01). Specifically, concentrations were consistently higher in females than males across all conditions. Comparing the interventions, cortisol levels at the end of the 8-hour heat event were higher in the Fan condition compared to both Con and Fan+Fluid for male participants, with no significant difference observed between Con and Fan+Fluid. In contrast, for female participants, cortisol concentrations were lowest in the Fan+Fluid condition compared to both Con and Fan at the end of the 8-h heat event.

### Perceptions

Perceptual responses to the 8-hour replicated heat event revealed significant improvements in thermal comfort, thermal sensation, thirst sensation, and psychological stress in the Fan+Fluid trial compared to both the Con and Fan trials (all *p*<0.05). However, no significant differences in these perceptual markers were observed between sexes across trials (all *p*>0.10). Specifically, participants in the Fan+Fluid trial reported greater thermal comfort, perceived less environmental heat, experienced reduced thirst, and noted lower psychological stress at the end of the heat event compared to those in the Con and Fan trials (**Table 2**).

**Table 2.**
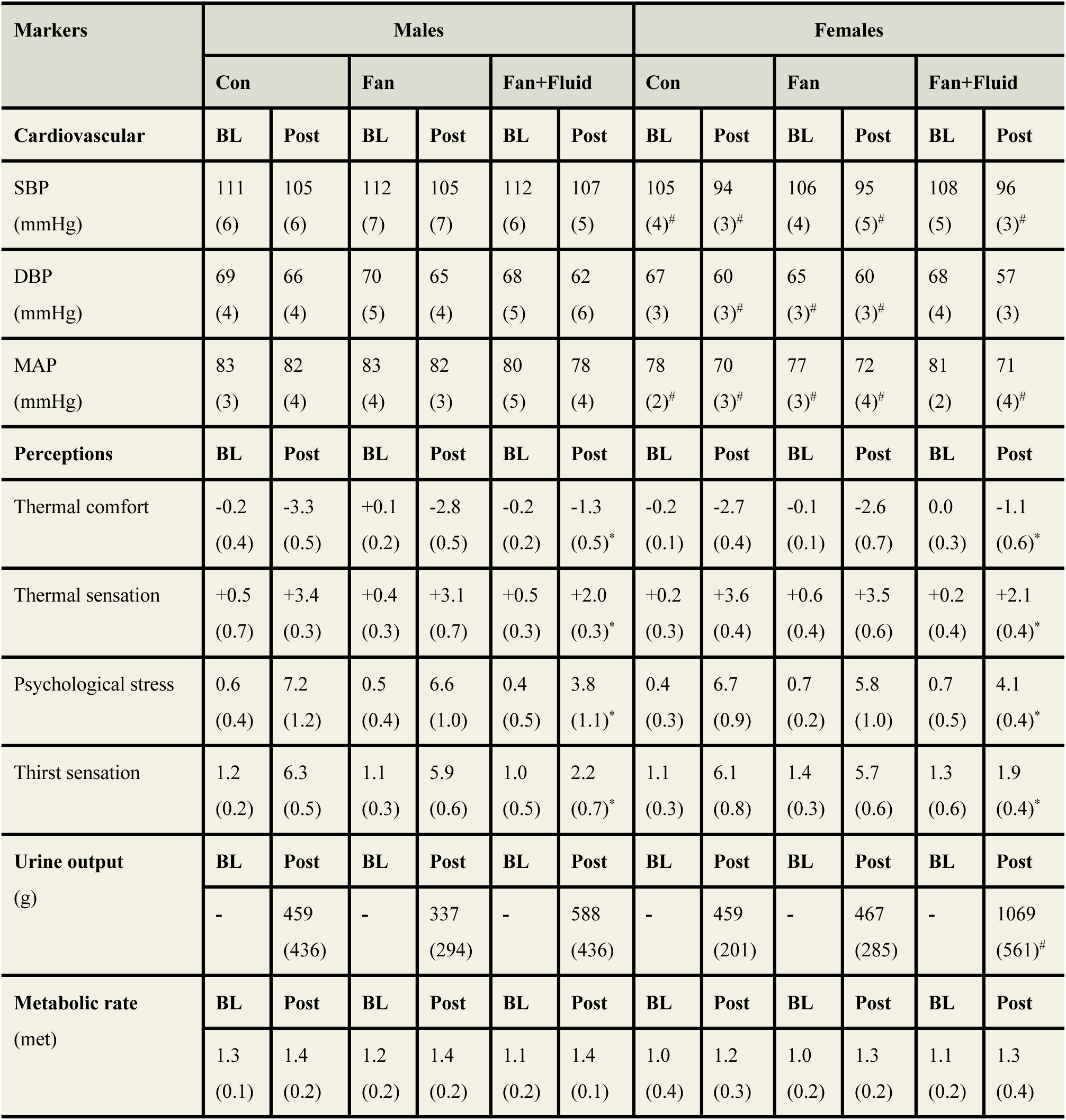
Cardiovascular, perceptual, urinary and metabolic markers measured at baseline (BL) and at the end (Post) of the 8-hour replicated August 3, 2024 Hangzhou heat event across conditions for male and female participants. -, not applicable; SBP, systolic blood pressure; DBP, diastolic blood pressure; MAP, mean arterial pressure; *, indicates Fan+Fluid significantly different from Con and Fan (*p*<0.05); #, indicates significant sex differences within the corresponding condition (*p*<0.05). +, indicates Fan+Fluid significantly different from Con (*p*<0.05). The numbers in parentheses represent standard deviation (SD).

| Markers | Males |  |  |  |  |  | Females |  |  |  |  |  |
| --- | --- | --- | --- | --- | --- | --- | --- | --- | --- | --- | --- | --- |
|  | Con |  | Fan |  | Fan+Fluid |  | Con |  | Fan |  | Fan+Fluid |  |
| Cardiovascular | BL | Post | BL | Post | BL | Post | BL | Post | BL | Post | BL | Post |
| SBP (mmHg) | 111<br>(6) | 105<br>(6) | 112<br>(7) | 105<br>(7) | 112<br>(6) | 107<br>(5) | 105<br>(4)<sup>#</sup> | 94<br>(3)<sup>#</sup> | 106<br>(4) | 95<br>(5)<sup>#</sup> | 108<br>(5) | 96<br>(3)<sup>#</sup> |
| DBP (mmHg) | 69<br>(4) | 66<br>(4) | 70<br>(5) | 65<br>(4) | 68<br>(5) | 62<br>(6) | 67<br>(3) | 60<br>(3)<sup>#</sup> | 65<br>(3)<sup>#</sup> | 60<br>(3)<sup>#</sup> | 68<br>(4) | 57<br>(3) |
| MAP (mmHg) | 83<br>(3) | 82<br>(4) | 83<br>(4) | 82<br>(3) | 80<br>(5) | 78<br>(4) | 78<br>(2)<sup>#</sup> | 70<br>(3)<sup>#</sup> | 77<br>(3)<sup>#</sup> | 72<br>(4)<sup>#</sup> | 81<br>(2) | 71<br>(4)<sup>#</sup> |
| Perceptions | BL | Post | BL | Post | BL | Post | BL | Post | BL | Post | BL | Post |
| Thermal comfort | -0.2<br>(0.4) | -3.3<br>(0.5) | +0.1<br>(0.2) | -2.8<br>(0.5) | -0.2<br>(0.2) | -1.3<br>(0.5)<sup>*</sup> | -0.2<br>(0.1) | -2.7<br>(0.4) | -0.1<br>(0.1) | -2.6<br>(0.7) | 0.0<br>(0.3) | -1.1<br>(0.6)<sup>*</sup> |
| Thermal sensation | +0.5<br>(0.7) | +3.4<br>(0.3) | +0.4<br>(0.3) | +3.1<br>(0.7) | +0.5<br>(0.3) | +2.0<br>(0.3)<sup>*</sup> | +0.2<br>(0.3) | +3.6<br>(0.4) | +0.6<br>(0.4) | +3.5<br>(0.6) | +0.2<br>(0.4) | +2.1<br>(0.4)<sup>*</sup> |
| Psychological stress | 0.6<br>(0.4) | 7.2<br>(1.2) | 0.5<br>(0.4) | 6.6<br>(1.0) | 0.4<br>(0.5) | 3.8<br>(1.1)<sup>*</sup> | 0.4<br>(0.3) | 6.7<br>(0.9) | 0.7<br>(0.2) | 5.8<br>(1.0) | 0.7<br>(0.5) | 4.1<br>(0.4)<sup>*</sup> |
| Thirst sensation | 1.2<br>(0.2) | 6.3<br>(0.5) | 1.1<br>(0.3) | 5.9<br>(0.6) | 1.0<br>(0.5) | 2.2<br>(0.7)<sup>*</sup> | 1.1<br>(0.3) | 6.1<br>(0.8) | 1.4<br>(0.3) | 5.7<br>(0.6) | 1.3<br>(0.6) | 1.9<br>(0.4)<sup>*</sup> |
| Urine output (g) | BL | Post | BL | Post | BL | Post | BL | Post | BL | Post | BL | Post |
|  | - | 459<br>(436) | - | 337<br>(294) | - | 588<br>(436) | - | 459<br>(201) | - | 467<br>(285) | - | 1069<br>(561)<sup>#</sup> |
| Metabolic rate (met) | BL | Post | BL | Post | BL | Post | BL | Post | BL | Post | BL | Post |
|  | 1.3<br>(0.1) | 1.4<br>(0.2) | 1.2<br>(0.2) | 1.4<br>(0.2) | 1.1<br>(0.2) | 1.4<br>(0.1) | 1.0<br>(0.4) | 1.2<br>(0.3) | 1.0<br>(0.2) | 1.3<br>(0.2) | 1.1<br>(0.2) | 1.3<br>(0.4) |

## DISCUSSION

Our findings demonstrate that electric fan use, when combined with adequate fluid replacement (3L), during prolonged extreme heat exposure. This challenges the conventional reliance on core temperature (*T_core_*) as the primary metric for evaluating fan efficacy. While prior studies have suggested that electric fans offer limited benefits above 33°C,^11,19–21^ our results establish that electric fans can suppress inflammatory responses during extreme heat, but only when fluid intake is sufficient.

The observed sex differences in inflammatory and organ function biomarkers likely stem from distinct physiological and hormonal mechanisms. Estrogen is known to amplify cytokine responses and influence thermoregulatory, though the precise pathways remain insufficiently explored.^34^ Our data indicate that females exhibited significantly higher IL-6, IL-1β, and IFN-γ levels than males across all three trial conditions, suggesting a heightened inflammatory response. Given that our study controlled for menstrual cycle phase by testing females during the early follicular phase (when progesterone is low), the potential influence of other hormonal fluctuations, such as estrogen peaks and inter-individual variability in menstrual cycle dynamics, remains unexplored. Previous studies have suggested that menstrual cycle phases may affect sweat rate, cardiovascular responses, and thermal perception,^35^ all of which could influence susceptibility to heat stress. Future studies should incorporate hormonal profiling and assess multiple cycle phases to clarify their role in thermoregulatory differences.

Our findings highlight the need for sex-specific interventions during extreme heat events. Broadening health assessments beyond core temperature to include systemic inflammation and organ function markers is critical, as these metrics directly influence the progression of heat-related illnesses. Rehydration strategies are particularly important for vulnerable populations in regions experiencing severe heat stress, such as southern and eastern China, the Persian Gulf, and the Red Sea, where environmental conditions similar to the Hangzhou heat event (August 3rd, 2024) are prevalent.^25,36–38^

Electric fan use, when paired with sufficient fluid replenishment, effectively suppresses inflammatory markers (23.9% reduction in IL-6 and 32.5% reduction in IL-1β) and organ function markers (15.9% reduction in ALT, and 23.6% reduction in BUN) compared to the control condition for both sexes. Conversely, electric fan use with limited fluid intake exacerbates inflammatory responses (e.g., IL-6: 35.7% higher in Fan vs. Con, 78.1% higher in Fan vs. Fan+Fluid) and organ dysfunction (ALT: 19.7% higher in Fan vs. Con, 42.0% higher in Fan vs. Fan+Fluid) for both sexes, likely due to increased dehydration and stress hormone release (**Table 1**). These findings are particularly relevant for informing public health recommendations to reduce heat-related illnesses.

Our study is the first to provide clinically relevant data partially supporting the WHO guideline on electric fan use during extreme heat events.^7^ While the WHO recommends fan use at temperatures up to 40°C, our study encompassed a broader temperature range (37.5–41.4°C), demonstrating fan efficacy even beyond the established threshold. This suggests that the current WHO guideline may be overly conservative for young adults under controlled hydration and warrants further evaluation, particularly under real-world conditions with dynamic environmental fluctuations. In contrast, our findings are inconsistent with the CDC guideline, which recommends a much lower threshold of 32.2°C,^6^ and a recent review by Meade et al., which suggests 35°C as the cutoff for electric fan use.^8^ This discrepancy arises from the reliance on a single health metric (i.e., core temperature) to determine electric fan efficacy while neglecting critical inflammatory and organ function biomarkers.^11,19–21^ Core temperature alone is a poor predictor of heatstroke onset^16^ and must be considered alongside inflammatory markers to identify systemic inflammatory response syndrome (SIRS), a key condition that elevates heatstroke risk.^39^ We urge the CDC to revise its guideline, as our study demonstrates that that electric fan use under the conditions of this replicated heat event effectively reduces heat-related illness risk, particularly for young adults. Additionally, we advocate for a broader evaluation framework that incorporates inflammatory responses and organ function markers to assess cooling interventions.

Furthermore, electric fan use during humid heat events should be tailored based on sex differences in physiological responses. Females may require stronger cooling strategies than males to reduce cardiovascular strain and prevent the progression from an elevated inflammatory response to SIRS, a critical condition that increases heatstroke risk. This is supported by our observation that heart rate (HR) did not decrease in the Fan+Fluid condition (**Figure 2**), and that inflammatory marker increases, such as IL-6 (Con: 257.9% higher; Fan: 212.2% higher; Fan+Fluid: 250% higher), were more pronounced in females than males by the end of the 8-hour heat exposure (**Table 1**).

While our results confirm that electric fan use effectively lowers inflammatory response during the replicated heat event in Hangzhou, with an average wet-bulb temperature: of 30.0°C (*T_W_*=29.1-30.7°C), our previous research has shown that electric fan use with adequate fluid intake becomes ineffective at reducing inflammation when the wet-bulb temperature rises to 32.0°C (ambient temperature: 40°C, 55% RH).^39^ This provides new insights into guidelines for electric fan use during humid heat events. Specifically, based on current and previous findings, electric fan use should be encouraged when wet-bulb temperature is ≤30.7°C, as it significantly reduces IL-6, IL-1β, ALT, BUN, and IFABP levels by 23.9%, 32.5%, 15.9%, 23.6%, and 64.7%, respectively, compared to the control. However, when the wet-bulb temperature reaches 32.0°C, alternative cooling strategies may be necessary, as fan use alone fails to suppress inflammatory responses.

Current health agency guidelines and prior research have primarily used dry-bulb temperature without specifying relative humidity, which can lead to inconsistent recommendations.^8^ Many studies have assessed fan efficacy at limited temperature-humidity points, producing conflicting results. Our data suggest that wet-bulb temperature (*T_W_*) should be used as a more precise threshold for electric fan use, as it integrates both temperature and humidity into a single physiologically relevant metric. Based on our findings, a *T_W_* of ≤30.7°C encourages fan use, whereas *T_W_* ≥32.0°C discourages it. However, the effects of fan use in the intermediate range (*T_W_*=30.7–32.0°C) remain unclear and require further investigation. We recommend that future guidelines incorporate wet-bulb temperature thresholds rather than relying solely on dry-bulb temperature to provide clearer and more applicable recommendations for fan use during extreme heat events.

## LIMITATIONS

This study has several limitations that warrants further investigation. First, the replicated heat event examined the health efficacy of electric fan use only during acute exposure, but did not assess its effectiveness during consecutive days of heat wave exposure, which is more representative of real-world conditions. Second, we have not identified the most effective mitigation strategy for females during replicated heat events. Given that females may require more intense cooling interventions than males to suppress their elevated inflammatory response and prevent heat-related illnesses, future studies should focus on sex-specific cooling strategies for prolonged heat exposure, rather than adopting a one-size-fits-all approach.

Moreover, our study did not account for real-world conditions where multiple cooling strategies are often used simultaneously. Electric fans are frequently combined with air conditioning, cooling vests, or intermittent water spraying, which may further enhance thermoregulation and mitigate heat strain. Assessing the synergistic effects of these interventions will be critical for refining public health recommendations and ensuring optimal cooling strategies, particularly for females who demonstrate heightened inflammatory and organ stress responses under extreme heat. Additionally, while our findings indicate that fan use is beneficial at *T_W_* ≤30.7°C and ineffective at *T_W_* =32.0°C, we did not examine its impact within the 30.7–32.0°C *T_W_* range, leaving a gap in knowledge that future studies should address. Research should evaluate the combined effects of fan use with other passive and active cooling methods and investigate the effectiveness of fan use in this intermediate *T_W_* range to develop targeted, evidence-based cooling strategies that maximize heat tolerance across sexes.

## CONCLUSIONS

Taken together, our study provides robust evidence supporting the use of electric fans during prolonged heat events under environmental conditions similar to the Hangzhou heat event. Our findings challenge existing guidelines that rely solely on core temperature thresholds to evaluate the efficacy of fan use. By demonstrating the significant reductions in systemic inflammation (IL-6 and IL-1β by 23.9% and 32.5%, respectively) and organ function markers (ALT and BUN decreased by 15.9% and 23.6%, respectively) when fans are combined with adequate fluid intake, this study highlights the critical importance of integrating inflammatory and organ function biomarkers into the evaluation of heat mitigation strategies. Moreover, the observed sex differences in inflammatory responses and cardiovascular strain underscore the necessity of tailoring heat mitigation strategies to specific populations, particularly females who may require more intensive cooling interventions to mitigate their heightened susceptibility to systemic inflammatory responses.

These insights not only validate the WHO recommendation on fan use but also call for a revision of current CDC guidelines to reflect a broader and more comprehensive set of health metrics beyond core temperature alone. Furthermore, our findings offer actionable guidance for regions prone to extreme heat events, such as southern and eastern China, the Persian Gulf, and the Arabian Gulf, where similar or more severe environmental conditions prevail. By encouraging the use of electric fans paired with fluid replenishment under specific thermal conditions, this study provides a practical, accessible and low-carbon strategy to reduce the risk of heat-related illnesses. Future research should focus on optimizing cooling interventions for females and assessing the cumulative effects of fan use over consecutive heatwave days, and determine the wet-bulb temperature upper limit at which fan use remains effective to further enhance public health outcomes.

## Data Availability

All data produced in the present study are available upon reasonable request to the authors

## Data availability statement

The data supporting the present findings are available from the corresponding author upon reasonable request. Due to privacy or ethical restrictions, the data is not publicly available.

## Acknowledgements

This research was financially supported by the National Excellent Young Scientist Program (grant number: 6119924022, to FW). We would also like to express our gratitude to the participants who took part in the study for their invaluable contributions to this research.

## Author contributions

All authors from this study meet the International Committee of Medical Journal Editors (ICMJE) criteria for authorship, and all those who qualify are listed. All authors had full access to and accept responsibility for the data presented in the study. FW conceived the research question and acquired funding. FW designed the trial. FW, TL, HW, and HX collected data. FW, HW, TL, and HX processed data. TL performed statistical analysis and created the data visualizations. FW drafted the manuscript. All authors revised the manuscript for important intellectual content.

