## Supplemental Table 1 for "Electric Fan Use in Replicated 8-Hour Extreme Heat Event: Sex Differences in Systemic Biomarkers and Thermoregulation" (from another version of this preprint: version 1, 2025-03-07)

### Supplemental Material

**Table S1** The start time, selected air speeds, and total fanning time at high air speed (AS) during the 8-hour extreme heat event. Data are stratified by sex to highlight differences in electric fan usage patterns among male and female participants.

| | Participants | Start time (seconds) | AS selection | Total fanning time (min) |
|---|---|---|---|---|
| **Males** | M1 | 1 | High | 480 |
| | M2 | 1 | High | 480 |
| | M3 | 1 | High | 480 |
| | M4 | 1 | High | 480 |
| | M5 | 1 | High | 480 |
| | M6 | 1 | High | 480 |
| | M7 | 1 | High | 480 |
| | M8 | 1 | High | 480 |
| | M9 | 1 | High | 480 |
| | M10 | 1 | High | 480 |
| **Females** | F1 | 1 | High | 480 |
| | F2 | 1 | High | 480 |
| | F3 | 1 | High | 480 |
| | F4 | 1 | High | 480 |
| | F5 | 1 | High | 480 |
| | F6 | 1 | High | 480 |
| | F7 | 1 | High | 480 |
| | F8 | 1 | High | 480 |
| | F9 | 1 | High | 480 |
| | F10 | 1 | High | 480 |

Note: M, males; F, females.
